# Development and assessment of the performance of a shared ventilatory system that uses clinically available components to individualize tidal volumes

**DOI:** 10.1101/2020.12.09.20246165

**Authors:** David M. Hannon, Tim Jones, Jack Conolly, Conor Judge, Talha Iqbal, Atif Shahzad, Michael Madden, Frank Kirrane, Peter Conneely, Brian H. Harte, Martin O’Halloran, John G. Laffey

## Abstract

**Objectives:** To develop and assess the performance of a system for shared ventilation that uses clinically available components to individualize tidal volumes under a variety of clinically relevant conditions.

**Design:** Evaluation and in vitro validation study.

**Setting:** Ventilator shortage during the SARS-CoV-2 global pandemic.

**Participants:** The design and validation team consisted of intensive care physicians, bioengineers, computer programmers, and representatives from the medtech sector.

**Methods:** Using standard clinical components, a system of shared ventilation consisting of two ventilatory limbs was assembled and connected to a single ventilator. Individual monitors for each circuit were developed using widely available equipment and open source software. System performance was determined under 2 sets of conditions. First, the effect of altering ventilator settings (Inspiratory Pressure, Respiratory rate, I:E ratio) on the tidal volumes delivered to each lung circuit was determined. Second, the impact of altering the compliance and resistance in one simulated lung circuit on the tidal volumes delivered to that lung and the second lung circuit was determined. All measurements at each setting were repeated three times to determine the variability in the system.

**Results:** The system permitted accurate and reproducible titration of tidal volumes to each ‘lung circuit’ over a wide range of ventilator settings and simulated lung conditions. Alteration of ventilator inspiratory pressures stepwise from 4-20cm H_2_O, of respiratory rates from 6-20 breaths/minute and I:E ratio from 1:1 to 1:4 resulted in near identical tidal volumes delivered under each set of conditions to each simulated ‘lung’. Stepwise alteration of compliance and resistance in one ‘test’ lung circuit resulted in reproducible alterations in tidal volume to the ‘test’ lung, with little change to tidal volumes in the ‘control’ lung (a change of only 6% is noted). All tidal volumes delivered were highly reproducible upon repetition.

**Conclusions:** We demonstrate the reliability of a simple shared ventilation system assembled using commonly available clinical components that allows individual titration of tidal volumes. This system may be useful as a temporary strategy of last resort where the numbers of patients requiring invasive mechanical ventilation exceeds supply of ventilators.

**Article Summary:** 

**Strengths and limitations of this study:**

- This solution provides the ability to safely and robustly ventilate two patients simultaneously while allowing differing tidal volumes in each limb.
- The designed solution uses equipment readily available in most hospitals.
- Accurate and reproducible titration of tidal volumes to each ‘lung’ was possible over a wide range of ventilator settings.
- Alteration of one simulated ‘lung’ conditions had minimal impact on the tidal volumes delivered to the unaffected lung
- The system relies on patients being sedated and paralysed.
- We have not yet tested this solution *in vivo*, on COVID-19 patients.

## Introduction

The global pandemic of coronavirus disease, COVID-19, caused by the SARS-CoV-2 virus began in late 2019 and is ongoing. At the time of writing, the disease continues to kill thousands of people each day. Therapy for this condition is, at present, supportive although many trials for both specific therapeutic interventions and vaccines are ongoing^[1, 2]^. The pandemic has placed unprecedented pressures on Intensive Care Units (ICUs) throughout the world. Severe pulmonary issues seem to complicate approximately 15% of cases of infection^[3]^, and reports of the percentage of hospitalised COVID-19 patients who will require admission to an Intensive Care Unit (ICU) can reach as high as 20%^[4, 5]^, and approximately half of these patients are likely to require mechanical ventilation. The resulting pressure on resources has led to difficult ethical decisions regarding resource allocation^[6]^.

One solution to this issue, in a emergent setting, is to ventilate more than one patient from a single ventilatory source. This is not an idea that is unique to the COVID-19 pandemic, and has been the subject of equipment tests^[7, 8]^, testing in animals^[9]^, computer simulation^[10, 11]^, and has even been utilised in a clinical setting in limited numbers during extreme conditions^[12]^. Ventilator sharing is clearly a strategy of last resort, though reports of doctors being obliged to choose which patients should receive ventilatory support, and the possibility of similar scenarios appearing during future pandemics, underline the need to consider this strategy as part of a response that increases short term ventilatory capacity.

A limitation to many previously proposed systems of shared ventilation is inability of the system to provide individualised tidal volumes to each limb of the circuit. The share of the overall tidal volume delivered to each patient varies depending on circuit characteristics and the patient related conditions. Changes in the condition of one patient may therefore impact on the ventilation of the other patient. Most COVID-19 patients admitted to an ICU fulfil criteria for Acute Respiratory Distress Syndrome (ARDS)^[13]^, and key recommendations regarding the optimal respiratory management of patients with ARDS who are intubated and ventilated concern the implementation of a therapeutic strategy of ‘Lung Protective Ventilation’^[14]^, in which tidal volumes and pressures supplied to the lung by a ventilator are of a magnitude to minimise ‘Ventilator Induced Lung Injury (VILI)’. This involves providing mechanical ventilation to a ‘sick’ lung in order to effectively oxygenate the patient while avoiding further injury to that lung by subjecting it to pressures or volumes that will lead to overdistension and cause inflammatory damage^[15]^. In the context of shared ventilation, this requires a system in which each ventilatory limb of a shared circuit can adapt tidal volumes to allow for the safe ventilation of two patients.

Our objective in this study was to develop a simple but reliable system that permits shared ventilation using commonly available clinical components that allows individual titration of tidal volumes. The objective of this study is to evaluate our shared ventilation system through a range of clinically relevant conditions. Further information including videos of the use of the system can be accessed at www.galwayventshare.com.

## Methods

### Setting

The system was tested using an Intensive Care ventilator attached to the shared circuit, each of which ventilated a test lung. The tests were performed in a laboratory setting and were using testing equipment only.

### Components and Assembly

The system was assembled as per the schematic design in Figure 01. A full list of components is available in Appendix A, as are instructions for assembly. The ventilator used was a Puritan Bennet 980 model (Medtronic plc). The ventilator operated in Pressure Control mode throughout the tests. Each limb was connected to a Michigan Instruments’ Training and Test Lung (Michigan Instruments Inc). Data regarding tidal volumes and pressures was captured with a mixture of the Citrex H4 gas flow analyser (IMT Analytics AG) and a custom developed open-source solution developed using a custom pressure sensor processed through the open-source Python programming language. The monitoring system consists of a gas flow sensor (Sensirion, SFM3200/3300), a pressure sensor (Analog Microelectronics, AMS-5915-0200-D-B), USB serial sensor cable (Nicolay GmbH), a processing and display unit (Raspberry Pi, 7-inch standard screen), and software (open source creative commons license).

**Figure 01.**
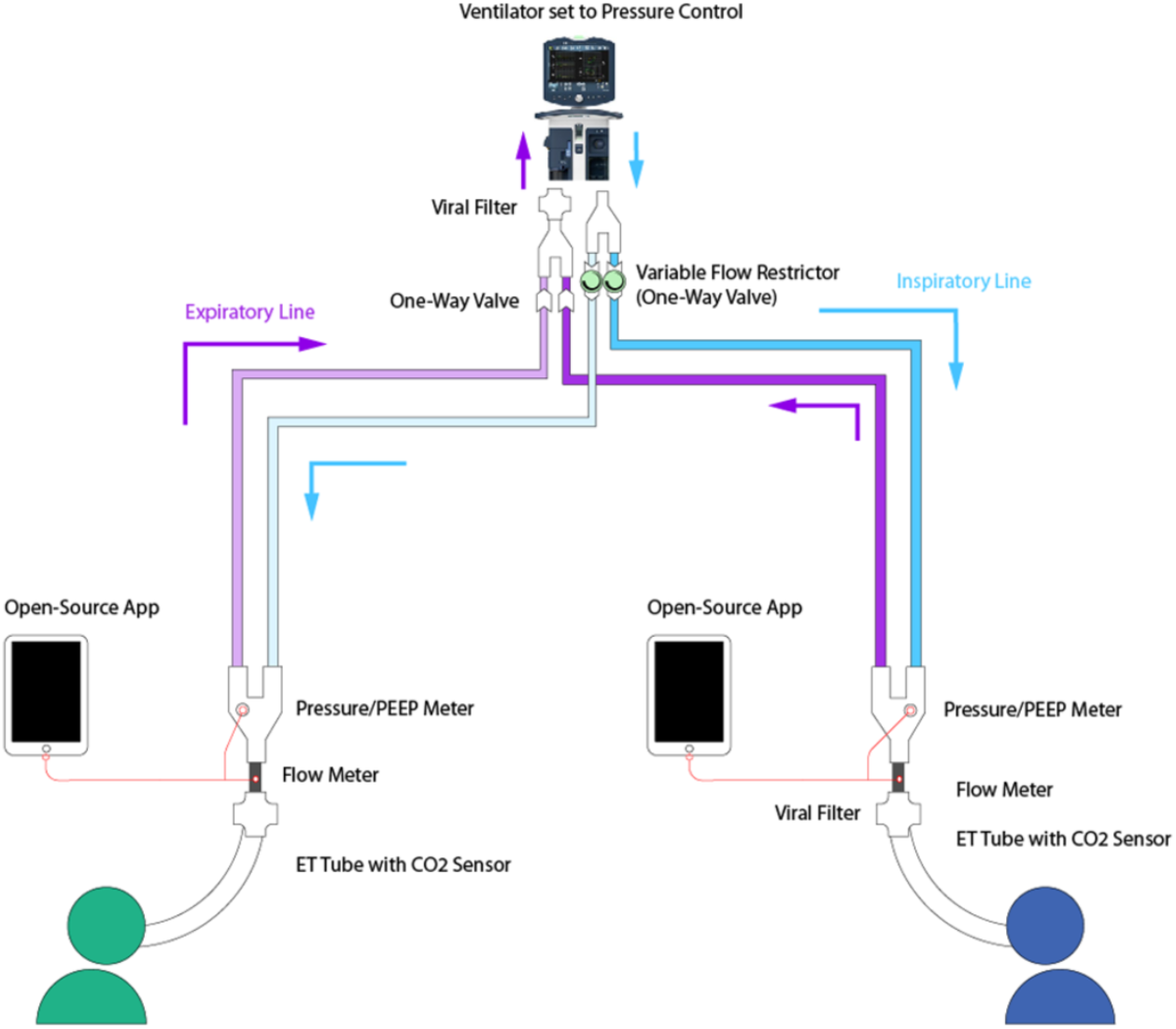
Diagram of the assembled shared ventilation system. Further details of the exact components used, and their assembly, are available in Appendix A.

Images of the assembled system and the patient monitoring output display are shown in Figures 02.

**Figure 02.**
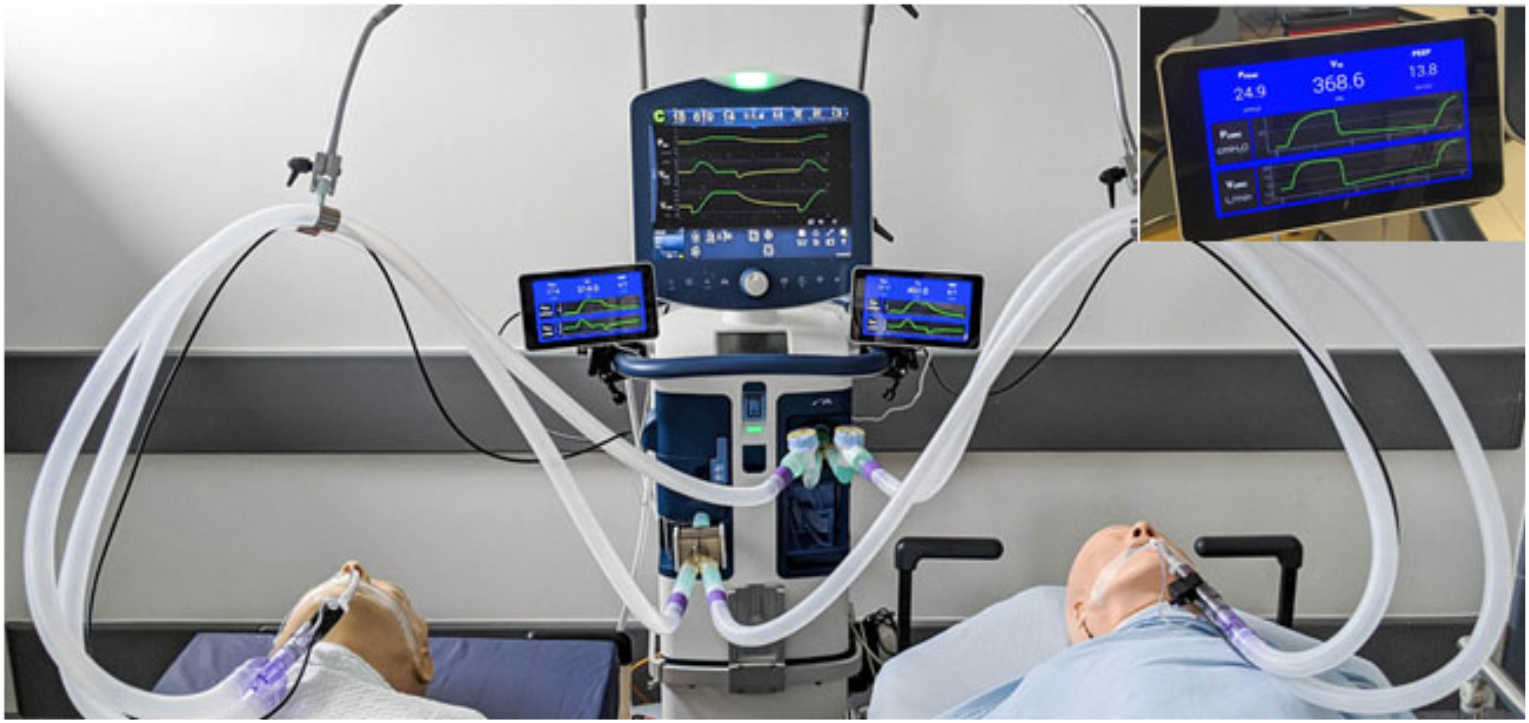
Image of the assembled system attached to two mannikins. Monitoring of the respiratory parameters for each limb are shown to the right and left of the main ventilator. A closeup of the display readout is also shown.

To assist with accuracy in testing the positions of the Adjustable Pressure-Limiting (APL) valves on each limb, plastic covers were added to the top of the valves. These covers were marked with ten equidistant dots to allow a more precise description of the position of the APL valve in each scenario. Position 0 corresponded to fully open (i.e. no resistance to flow), and position 10 to fully closed (i.e. near complete resistance to flow). These devices were developed by Design Partners, a Dublin-based design firm.

### Procedure

The system was subjected to repeated tests. These test runs fell into two main groups. In the first group of tests, a focus was placed on altering the settings of the ventilator and recording the effect of these changes on the tidal volumes delivered to each test lung. In the second group of tests, the system performance under different conditions in each limb of the shared circuit was recorded. A test was performed in which the APL setting of Limb A was manipulated, followed by a test in which the compliance of the test lung of Limb A was altered. Each of the aforementioned tests were repeated three times.

In addition to recording tidal volumes, the Peak End Expiratory Pressure and the Peak Pressure in each limb were recorded. The limbs of the circuit will henceforth be referred to as ‘Limb A’ and ‘Limb B’.

In the initial group of tests, the compliance of the test lung for Limb A and Limb B was 0.1L/cmH2O. The APL valve on each limb was set to mark 3 of 10. These values were unchanged throughout each run of the various tests. Standard ventilator settings were a respiratory rate (RR) of 14 breaths per minute, peak end expiratory pressure (PEEP) was 8 cmH2O, inspiratory pressure for each breath was 10cmH2O, and the I:E ratio was 1:2. In this initial group, a focus was placed on manipulating the aforementioned ventilator settings throughout a range of values that may be encountered clinically.

A series of three runs was performed in which the inspiratory pressure was manipulated through a range from 5 to 20cmH2O per breath, a series in which the RR was tested in increments between 6 and 20 breaths per minute, and a sequence of tests in which the I:E ratio was altered in increments from 1:1 to 1:4.

In the second group of experiments, in which effects to changes of Limb A were recorded on the system, the ventilator settings were set as follows; inspiratory pressure was set at 10 cmH2O, RR was 14 breaths per minute, PEEP was 5 cmH2O, and I:E ratio was 1:2. Unless otherwise altered, standard settings for each limb was a compliance of 0.1 L/cmH2O.

In the first run of experiments in this grouping, the compliance of Limb A was altered in a stepwise fashion from 0.01L/cmH2O to 0.15L/cmH2O. The APL valve of Limb A and Limb B was set to 3 of 10. In the second run of experiments in this group, lung compliance for each limb was set to 0.1L/cmH2O. The APL of Limb B was fully open (to ensure to pressure warnings were encountered from the ventilator), and the APL of Limb A was manipulated in a series of steps from 0 of 10 (fully open) to 10 of 10 (fully closed). As the APL valve closes, it increases resistance to gas flow through the APL in that limb. A setting of 0 is fully open (i.e. no extra resistance to gas flow) while full closure means that there is a maximal resistance to gas flow at this point.

## Results

### Alteration of ventilator settings

The results of the experiment wherein the settings for each limb are identical but the Inspiratory Pressure from the ventilator is titrated upwards show a linear increase in tidal volumes with very little discrepancy between each limb, and a small standard deviation at each setting. The results can be seen in Figure 03, panel A.

**Figure 03.**
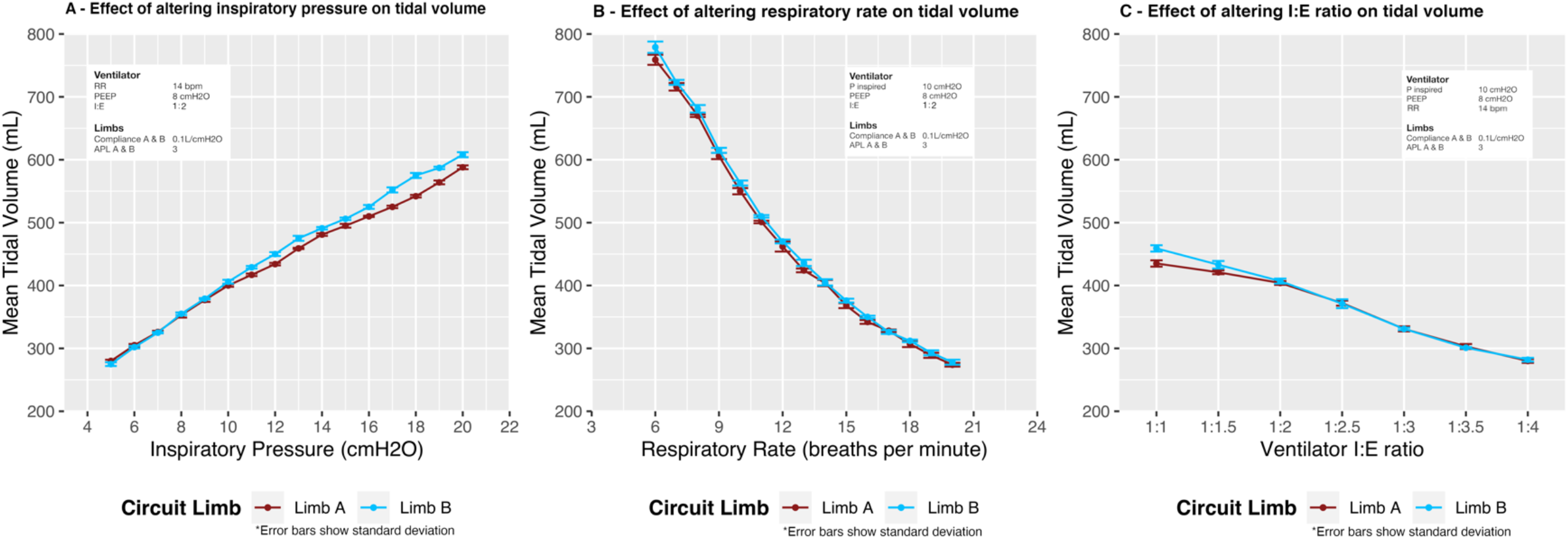
Effect of altering ventilatory parameters on tidal volumes delivered to simulated lungs ‘A’ and lung ‘B’. The tidal volumes delivered under each set of conditions to each simulated ‘lung’ were nearly identical and were highly reproducible upon repetition (n = 3 repetitions for each set of conditions)

Results from the test in which Respiratory Rate was increased show an expected inverse relationship between Respiratory Rate and Tidal Volume. Once again there is good agreement between the limbs, and a very narrow standard deviation. Details can be seen in Figure 04, panel B.

**Figure 04.**
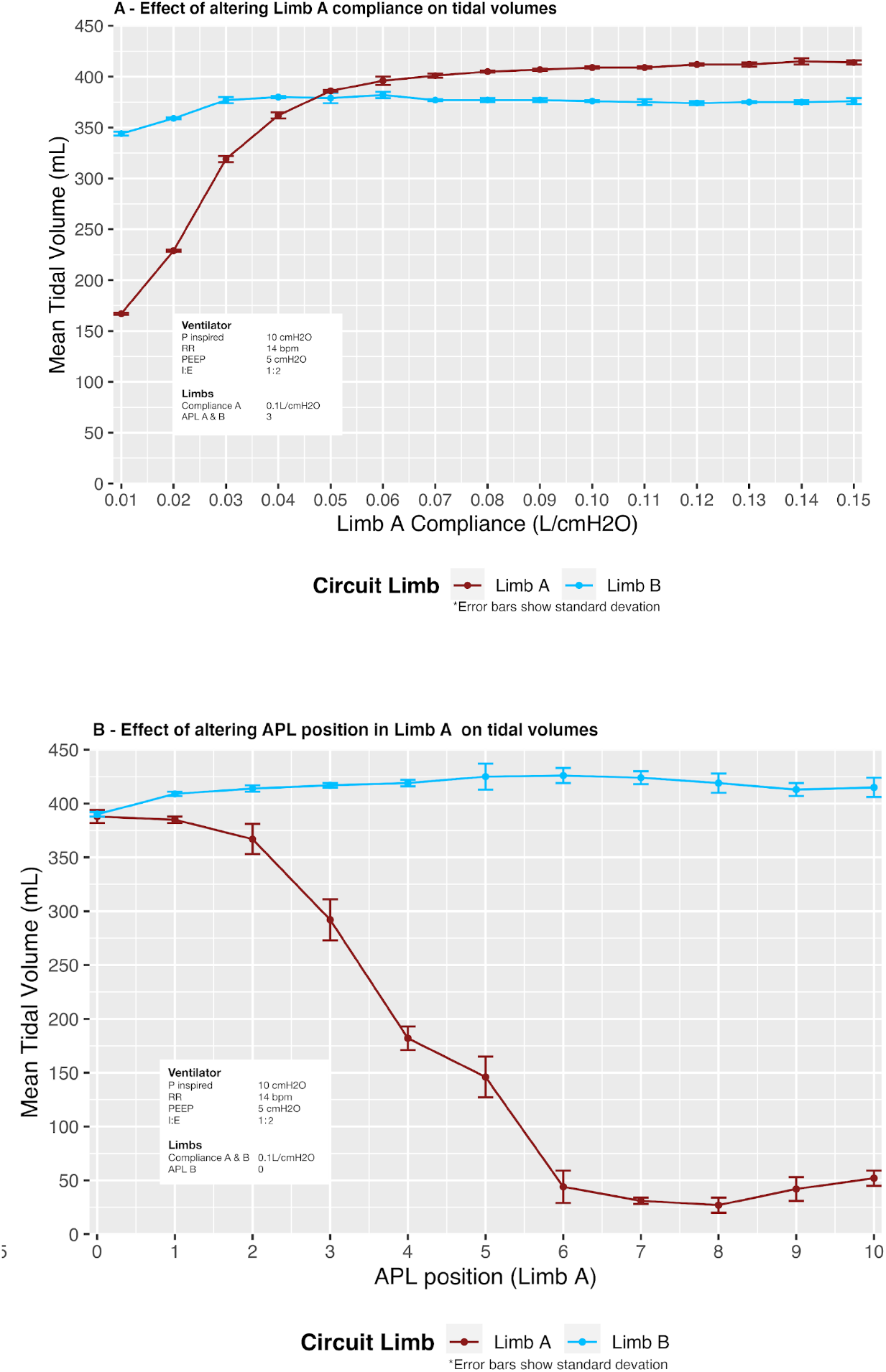
Effect of altering the conditions of one simulated ‘lung’ (Lung A) on tidal volumes delivered to both Lung A and Lung B. In panel A, the compliance of the test lung of Limb A was sequentially increased. In panel B, The APL valve of Limb B was fully open. The APL valve of Limb A was closed sequentially from 0 to 10, with tidal volumes recorded (n = 3 repetitions for each set of conditions). Closing the APL increases the resistance to gas flow in that limb only.

There was a small consist discrepancy in tidal volumes between Limb A and Limb B at an I:E ratio of 1:1 (24mL) which disappeared as the I:E ratio increased. As before, the limbs were in close agreement and standard deviation of the tidal volume values was low. Results can be seen in Figure 04, panel C.

Statistical analysis was performed, and graphics were generated, using the open-source statistical programming package R, version 4.0.2 (2020-06-22) - “Taking Off Again”.

### Alteration of limb settings

The experiment showing tidal volume alterations as the lung compliance is changed on Limb A show a consistent difference between Limb A and Limb B that does not change until lung compliance is reduced below 0.07 L/cmH2O. After this point, a marked reduction in the tidal volumes of Limb A was observed. Results are shown in Figure 05, panel A.

This experiment also showed that as the APL valve of Limb A is closed, a reduction in tidal volumes was observed with a small standard deviation calculated. A small rise in tidal volumes was observed as the APL valve on Limb A was closed beyond position 7. Details are shown in Figure 05, panel B. Detailed figures for this test are also visible in Table 01.

**Table 01.** Effect of altering APL valve on Limb A, and thereby raising resistance to gas flow in Limb A on tidal volumes and peak inspiratory pressures of Limb A and B. The APL of Limb B is constant at mark 0, and the ventilator settings remain constant throughout (inspiratory pressure was set at 10 cmH2O, RR was 14 breaths per minute, PEEP was 5 cmH2O, and I:E ratio was 1:2. Unless otherwise altered, standard settings for each limb was a compliance of 0.1 L/cmH2O) *All values summarised as mean (SD)±(range)

|  | Limb A |  | Limb B |  |
| --- | --- | --- | --- | --- |
| APL (Limb A) | Tidal volume (mL) | PEEP (cmH2O) | Tidal volume (mL) | PEEP (cmH2O) |
| 0 | 388 (6) ± (382-392) | 10.7 (0.4) ± (10.4-11.1) | 390 (2) ± (389-392) | 10.4 (0.1) ± (10.3-10.5) |
| 1 | 385 (3) ± (383-388) | 10.4 (0.2) ± (10.2-10.6) | 409 (2) ± (407-411) | 10.2 (0.2) ± (10.0-10.3) |
| 2 | 367 (14) ± (354-381) | 10.1 (0.2) ± (9.9-10.3) | 414 (3) ± (412-417) | 10.0 (0.2) ± (9.8-10.2) |
| 3 | 292 (19) ± (278-313) | 9.6 (0.5) ± (9.2-10.1) | 417 (2) ± (416-420) | 10.0 (0.2) ± (9.8-10.2) |
| 4 | 182 (11) ± (172-194) | 9.7 (0.2) ± (9.6-10.0) | 419 (3) ± (415-421) | 10.2 (0.0) ± (10.2-10.2) |
| 5 | 146 (19) ± (129-166) | 8.0 (2.1) ± (5.6-9.3) | 425 (12) ± (412-434) | 9.7 (0.5) ± (9.3-10.3) |
| 6 | 44 (15) ± (33-61) | 4.6 (0.8) ± (3.7-5.1) | 426 (7) ± (420-433) | 10.1 (0.4) ± (9.7-10.4) |
| 7 | 31 (3) ± (28-33) | 4.2 (1.1) ± (3.2-5.4) | 424 (6) ± (418-429) | 10.3 (0.2) ± (10.0-10.4) |
| 8 | 27 (7) ± (22-35) | 3.6 (0.2) ± (3.5-3.9) | 419 (9) ± (411-428) | 10.2 (0.3) ± (10.0-10.5) |
| 9 | 42 (11) ± (30-52) | 2.2 (0.3) ± (1.9-2.4) | 413 (6) ± (408-420) | 10.4 (0.2) ± (10.3-10.7) |
| 10 | 52 (7) ± (46-60) | 2.1 (0.2) ± (1.9-2.3) | 415 (9) ± (406-424) | 10.5 (0.3) ± (10.2-10.7) |

## Discussion

We have successfully constructed and tested a system that enables shared ventilation that allows for titration of tidal volumes independently in each limb through the innovative use of a paediatric APL valve.

The system clearly shows that changes in resistance to gas flow, and pressure in one limb have a minimal effect on the other. The use of one-way valves and filters ensure that each patient never breathes the same air as the other. The system allows tidal volumes that are titratable over a wide range of tidal volumes and is stable for long periods. Given the heterogenous nature of the lung mechanics involved in ventilated COVID-19 patient with ARDS^[16]^, the ability to titrate tidal volumes is key in ensuring that shared ventilation can be performed while limiting traumatic damage to already compromised lung tissue^[17]^.

Testing outlined in this paper took place using the aforementioned sophisticated critical care ventilator, but the system has also been tested on a broad range of ventilators and can be utilised with any ventilatory source that is capable of Pressure Control Ventilation. As is common to many similar systems, the use of Volume Control Ventilation can result is unpredictable delivery of tidal volumes to each limb and therefore risks hyperinflation of areas of lung tissue^[18]^. This forms an obvious and important area for further research, as lung protective ventilation has been shown in both trials and reviews^[19]^ to reduce mortality in ARDS. Enabling a system of shared ventilation to safely and reliably operate in this mode is ideal.

Any system of shared ventilation allows for the ‘expansion’ of equipment stocks and enables life-preserving therapy to be delivered to multiple patients simultaneously. However, any such system comes with significant and important drawbacks. As noted earlier, these drawbacks have even led many noted and important medical organisations to advocate against the use of shared ventilation^[20]^. However, we feel that further exploration of this area is an important step in a ‘last resort’ scenario. It is also important to note that the technical drawbacks of this system, and the equipment it requires are often not the limiting factor in delivering ventilatory support to critically ill patients in resource-poor settings^[21]^. The key factor that limits ventilatory support in poor settings, and has been supported as the most important step, is the ability to delivery supplemental oxygen^[22]^. As such, shared ventilation may be better suited to settings which find themselves with the relevant expertise, but with equipment stocks that are being overwhelmed. Effective strategies in these challenged settings often focus on much more straightforward steps to halt the spread and severity of the disease before it leads to critical illness^[23]^.

Any discussion of shared ventilation in the context of a strategy of last resort to preserve life must also acknowledge the significant and difficult ethical implications inherent in this act. This strategy necessitates depriving a single patient of ‘standard-of-care’ treatment in order to seek to preserve the life of two patients^[24]^. Taking this action in the absence of robust data of the efficacy of shared ventilation for COVID-19 patients is extremely challenging given that one cannot be certain that it will confer benefit. Despite the attempts that have been made to aid this^[25]^, the initiation of shared ventilation is an ethically challenging act.

There are many limitations to this approach, and it has even led some organisations to issue joint statements against its use^[20]^. Employing this solution demands a high level of sophisticated knowledge, staff training, and usually comes with significant restriction regarding the ability to alter respiratory parameters for each patient^[18]^. As such, the authors do not endorse or advocate ventilator sharing as a normal course of therapy. Our aim is to provide details of a system that can be used as a strategy of last resort by medical professionals who find themselves in a situation in which shared ventilation must be initiated to preserve life. Considerable skill is required to operate a ventilator sharing system and the authors can take no responsibility or liability for any injury or harm caused as a result of the use of this ventilation setup.

Despite the important innovation presented here, patients must still share a majority of respiratory supports whilst sharing a ventilator. This includes identical respiratory rates, positive end expiratory pressures, and fraction of inspired oxygen. The authors are continuing to explore solutions to these issues, as they are important factors continuing to restrict the scope of patients that can be initiated on shared ventilation^[26]^. In addition to sharing certain respiratory settings, the system requires that both patients are fully ventilated, as there is no facility to allow differing spontaneous breaths between the limbs. This necessitates that both patients are sedated and paralysed. Paralysis is not helpful beyond short time-frames^[27]^ in a critical care patient, and the risk of undue muscular atrophy must be carefully considered when embarking on a course of care that necessitates paralysis. An ideal solution would allow the titration of as many respiratory parameters as possible, could respond to changes in lung mechanics of patients, and accommodate as wide a range of patients as possible, whilst facilitating lung protective ventilation.

Our solution shows a difference in tidal volumes when both limbs have open APLs and the our ventilatory circuit is applied. A typical and consistent difference of 5-10% has been observed during testing. In addition, this difference changes slightly over the range of Inspiratory Pressures delivered by the ventilator. We have observed that the limb featuring an APL valve in a semi-closed position typically shows an increase of 9-10mL per breath, versus 12-13mL if the APL is fully open. This is an important operational feature that operators would have to be aware of and also emphasises the need for detailed and repeated review of the circuit limbs if they were in use.

The monitoring system outlined above is a custom solution developed using commonly available parts from a number of online and regional local resellers worldwide. There are other systems available that seek to use the information obtained from an arterial pressure transducer to achieve these aims^[28]^, and this could enable an alternative solution to this issue than the one we present here.

Although the system remains stable over the majority of APL settings, a relative decrease in PEEP can be seen when the APL moves beyond point 5. This limits the utility of the system if it must operate with the APL in a position, as might be the case if the two patients who are sharing the system have very disparate tidal volume targets, or if there were significant changes in lung compliance. The loss of PEEP at these extremes can have important implications for patients with COVID-19, as the lungs of these patients are often highly responsive to PEEP^[29]^. If the two patients have been matched based on similar PEEP requirements, a large difference in this value could be detrimental, and emphasises the need for frequent patients monitoring.

High levels of skill and knowledge are typically needed to operate any system involving shared ventilation^[30]^. This, plus the importance of detailed monitoring as mentioned above could unduly add to the workload required when operating the system. The authors are both seeking to address this issue and are also conducting simulation studies to investigate the ability of clinicians to safely and effectively operate the system.

## Conclusions

We have showed that a system of shared ventilation using commonly available clinical components that allows titration of tidal volumes is possible and can be assembled easily. The detailed monitoring of ventilatory support delivered to each patient remains important and is now possible using cheap equipment and free open-source software.

Further work is necessary to determine ways in which additional basic respiratory parameters can be altered in each limb, but we believe our solution presents several important innovations.

In conclusion, we must still maintain the position that, in an ideal world, multiple patients would never be placed on a single ventilator. However, in an extreme scenario that requires the preservation of life we believe our solution offers an important innovation to increase the utility of this strategy of last resort.

## Data Availability

The data that support the findings of this study are available from the corresponding author, John G. Laffey, upon reasonable request.

## Contributors

DMH, TJ, JC, JL developed the Galway VentShare system. DMH, TJ, CJ, JL drafted the manuscript. AS, MM developed the software monitoring system for Galway VentShare. DMH, TJ, JC, CJ, FK, PC, BH, MOH, JL all reviewed manuscript prior to submission.

## Patient consent for publication

Not required.

## Appendix A System Assembly

**Figure A01.**
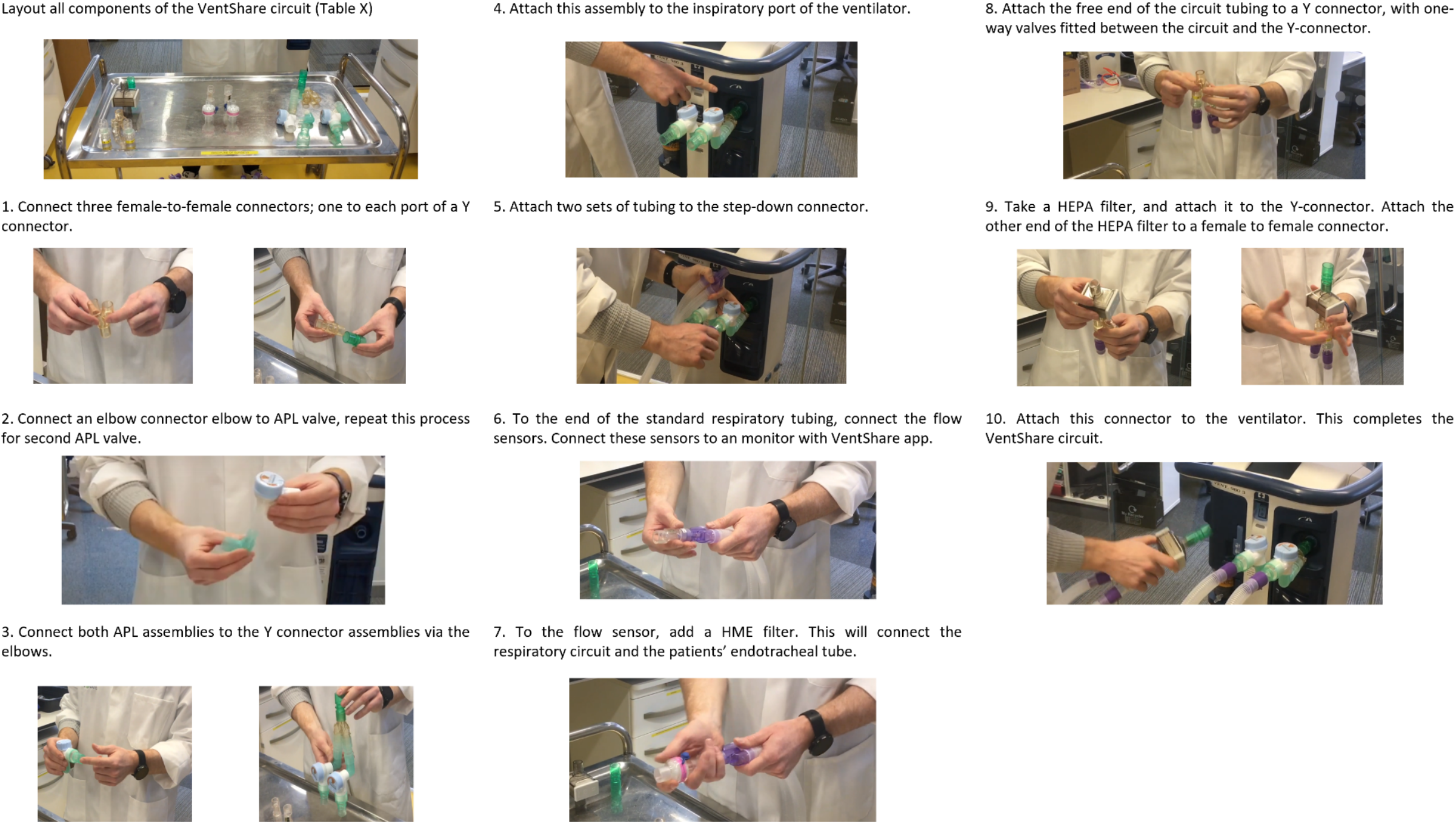
Stepwise assembly of ventilation system.

**Table A01.** Equipment list for the full assembly of the Galway VentShare system.

| Quantity | Item | Description |
| --- | --- | --- |
| 1 | Ventilator | An ICU ventilator offers the greatest control and sophistication, but the VentShare system has been tested on simpler ventilators and has shown comparable performance. |
| 2 | Ventilator circuit | Ensuring that these circuits are of sufficient length to reach each patient, but as short as possible within this constraint in order to minimise apparatus dead space. |
| 2 | Y-connector | 22mm male to 2 x 22mm male connectors |
| 2 | APL Valve | Paediatric APL valves from Mapleson anaesthetic circuits have been found to give reliable performance in testing. Importantly, these valves must incorporate a 30mm scavenging port. |
| 2 | Elbow connector | Connect directly to the APL valves and to the respiratory tubing. We have used 15mm male to 22mm male connectors. |
| 2 | Step-down connector | 30mm female to 22mm male connectors. |
| 4 | Female-to-female connector | Allows for connection of 22mm male to 22mm male connectors. |
| 2 | Flow sensors | The sensors used are manufactured by Sensirion. They are not necessary for operation of the circuit but are recommended to ensure accurate monitoring of each limb. |
| 2 | Heat and moisture exchange filter (HMEF) | We recommend HMEs that are capable of removing both bacteria and viruses (i.e. a dry circuit). |
| 1 | High Efficiency Particulate Air filter (HEPA filter) | This type of air filter can theoretically remove greater than 99.97% of any airborne particles with a size of 0.3 microns ($\mu\text{m}$). |
| 2 | One-way valves | Mounted on the expiratory limb, these are key to minimise 'cross-talk' between the individual limbs of the circuit. |
| 2 | Control Cap | Plastic caps covering the APL dials, providing more detailed visual feedback for the degree of APL closure. |

